# Plasma-treatment with the virucidal beta-propiolactone does not preclude analysis by large panel affinity proteomics, including the discovery of biomarker candidates

**DOI:** 10.1101/2023.06.29.23292027

**Authors:** Vanessa M. Beutgen, Aditya M. Bhagwat, Silke Reinartz, Rolf Müller, Johannes Graumann

## Abstract

Virus inactivation is a prerequisite for safe handling of high-risk infectious samples. Beta-propiolactone (BPL) is an established and commonly used reagent with proven virucidal efficacy. BPL primarily reacts with DNA and RNA, but also amino acids. The latter may yield modified antigenic protein epitopes and thus interfere with the binding properties of affinity reagents such as antibodies and aptamers, including panels of such reagents used in affinity proteomic screens. We investigated the impact of BPL-treatment on the analysis of protein levels in plasma samples using the commercial aptamer-based affinity proteomic platform SomaScan. Heparin-plasma samples from patients with ovarian cancer (n = 12) and benign tumors (n = 12) were analyzed using the SomaScan v4.1 platform, which led to the identification of COL10A1 as a novel ovarian cancer biomarker, a protein strongly associated with poor clinical outcome. BPL-related changes in protein detection were evaluated comparing native and BPL-treated state, simulating virus inactivation, and impact on measurable group differences was assessed. While approximately one third of protein measurements were significantly changed by the BPL treatment, a majority of antigen/aptamer interactions remained unaffected. Interaction effects of BPL treatment and disease state, potentially altering detectability of group differences, were observable for less than one percent of targets (0.6%). Accordingly, noticeable global effects of BPL treatment did not interfere with detectability of differential protein expression between benign and ovarian cancer samples, as measurements are altered in both groups to the same extent. Global effect sizes (Cohen’s d) between benign and cancer in BPL-treated samples and the number of significantly altered protein abundance observed in limma-based linear modeling appeared minimally increased, slightly enhancing the probability of false positive hits. Taken together, the results indicate the SomaScan platform as well suited for the analysis of high biosafety risk samples inactivated using BPL and the identification of novel biomarkers.

## Introduction

The investigation of viral diseases with the capacity to cause global pandemics or severe local outbreaks is a major challenge for medical research. Newly emerging OMICs technologies are well suited to gain deeper understanding of the associated pathophysiology and may open new treatment avenues. Blood and its derivatives provide highly informative samples that are easily accessed and subjected to comprehensive analysis. For downstream processing, blood products from patients with high-risk infectious viral disease are commonly treated with β-propiolactone (BPL) to inactivate the infectious agent. BPL-treated samples are considered non-infectious and efficacy was reported for a broad range of viruses [1]. Seeking an agent with virucidal efficacy paralleled by minimal effects on other blood components, Gerald LoGrippo in 1960 selected BPL from a collection of 23 virucides. BPL is unstable in aqueous solutions, as it gradually hydrolyses. Its half-life is highly temperature dependent with 3–4 h at room temperature, yielding the degradation products β-propionic acid and hydracrylic acid derivates [1], [2].

Studies investigating the effect of BPL on downstream assays have come to diverging conclusions. Ball et al. tested the effects of BPL on immunological analysis of various blood components of common interest in the context of HIV infection [3]. They tested for G, A and M class immunoglobulins, complement components C3 and C4, haptoglobin, α1-antitrypsin, C-reactive protein and others using a Beckman Immunochemistry Analysis System. Measured plasma concentrations of these proteins did not change significantly as a result of BPL treatment. They further tested for concentrations of various auto-antibodies and could not detect clinically relevant alterations either. Similar conclusions were derived from a second study that investigated the effect of BPL on antibody measurements by ELISA [4]. Here, serum levels of antibodies against tetanus toxin, diphtheria toxin, Haemophilus influenzae type b capsular polysaccharide and pneumococcal polysaccharide in BPL-treated and untreated samples from the same subjects were analyzed. Antibody concentrations in treated and untreated samples showed high correlation, yielding the conclusion that virus inactivation by BPL does not interfere with accuracy of measurements in ELISA, at least not for the concentrations tested. These findings, however, appear not generally reproducible or generalizable to all proteins. Although, α1-antitrypsin (AAT) levels were previously reported to be unchanged by BPL-treatment, this result was not reproduced in other work [5], which investigated the impact of sample pH, incubation time and temperature on apparent protein abundance. A decrease in measured AAT abundance was observed to correlate with pH, time and BPL concentration. Samples analyzed were treated with relatively high BPL concentrations and not buffered under all conditions tested. AAT levels were further found to continuously decrease with storage time following BPL incubation.

Taking together the findings of these studies, it may be hypothesized that low levels of BPL in a buffered sample do not influence detectability of a majority of blood proteins when storage time between treatment and measurement remains limited. BPL concentrations commonly used in virus inactivation protocols range from 0.025 to 1% [6]. Optimal concentrations depend on the target pathogen and need to be determined experimentally [7]. Lower concentrations may generally be desirable to reduce unwanted side effects, e.g. on proteins. More recently, the effects of BPL on different nucleobase analogues and amino acid residues were investigated systematically and a plethora of BPL-related protein modifications were reported [6], inviting speculation on the mechanisms of the observed impact of BPL on affinity/epitope interaction [8].

With the appearance of next-generation plasma and serum proteomics in biomarker discovery studies, a need arises for the investigation of the impact of BPL treatment on blood-derived samples in large-scale, high-throughput affinity proteomic analysis, as provided by the SomaScan platform. To validate the use of such technologies on BPL-treated samples, two main questions need to be answered: (1) Does the treatment affect the measurements obtained by the platform and (2) if so, to what extend does it impact on detection of differential protein expression between study groups? We set out to answer these questions and evaluated BPL-treatment compatibility with such platforms by analyzing the effect of low-concentration, bicarbonate-buffered BPL treatment on 50% diluted plasma samples of patients with benign tumors (n = 12) and ovarian cancer (n = 12) with respect to protein detectability in the aptamer-based proteomic platform SomaScan.

## Materials & Methods

### Plasma samples

Heparin-plasma samples were collected from patients with high-grade ovarian carcinoma (n=12) and patients with non-cancerous (benign) tumors (n=12). Samples were collected according to the guidelines of the Declaration of Helsinki with the informed consent of the patients and approval by the ethics committee of Marburg University (205/10). All patients agreed in writing to the publication of pseudonymized data derived from clinical material. Plasma was diluted 1:2 with PBS prior to storage at -80°C.

### β-propiolactone (BPL) treatment

Prior to analysis using the SomaScan platform, samples were buffered by adding sodium hydrogen carbonate (NaHCO_3_) to a final concentration of 45 mM and one sample each per subject was either mock or BPL treated. To that end, subsequent to fresh dilution of BPL with dH_2_O to yield a 50% stock solution, 0.3 μL of the solution was added to buffered samples (300 μL), resulting in a final concentration of 0.05% BPL. BPL and mock (dH_2_O) treated samples were incubated for 72 h at 4°C.

### SomaScan Assay

All samples were analyzed on the SomaScan v4.1 platform by SomaLogic Inc. (Boulder, CO, USA). SomaScan is an aptamer-based affinity proteomics platform that can analyze approximately 7,000 proteins in small sample volumes simultaneously [9]. The platform provides semi-quantitative protein readouts reported as *Relative Fluorescence Units* (RFU). Features receiving a quality flag after data normalization by SomaLogic were included in downstream analyzes probing possible effects of BPL treatment.

### Amino Acid Composition

Calculation of relative amino acid frequencies was carried out for the top 100 proteins with significantly (Bonferroni corrected p < 0.05) increased or decreased values after BPL treatment. Human proteome information for SomaScan v4.1 targets was downloaded in FASTA format from UniProt (13.10.2022). Swissprot-reviewed entries and canonical sequences (no isoforms) were considered exclusively, resulting in 6,388 relevant protein sequences. The retrieved sequence data was used to calculate reference values for expected amino acid frequencies normalized to 1 and compared to the observed frequencies of the top 100 proteins significantly affected by BPL.

### Survival associations

Overall survival data were retrieved from the PRECOG database [10] at https://precog.stanford.edu.

### Statistical Analysis

SomaScan data was analyzed in R (v4.2.3) and RStudio (v2023.03.1) using the ‘autonomics’ package (Bhagwat A, Hayat S, Graumann J. autonomics: Generifying and intuifying cross-platform omics analysis. DOI: 10.18129/B9.bioc.autonomics). The ‘autonomics’ package initially performs a log2 transformation of the RFU values during data import. All RFU presented in this study represent log2 transformed values.

## Results

Overall, only minor reductions in signal intensity were found following BPL treatment, indicating that global protein detectability using SomaScan remains unaffected (Figure 1A). The effect was lower than what was observed for protein abundance differences between cancer and benign samples in each condition (Figure 1B & C). This was also reflected in principal component analysis, where a unidirectional shift was observed upon BPL treatment (Figure 1D). However, separation between benign and cancer samples was preserved, as protein abundances shifted to the same extent for both groups and relative levels appeared to remain unaltered.

**Figure 1:**
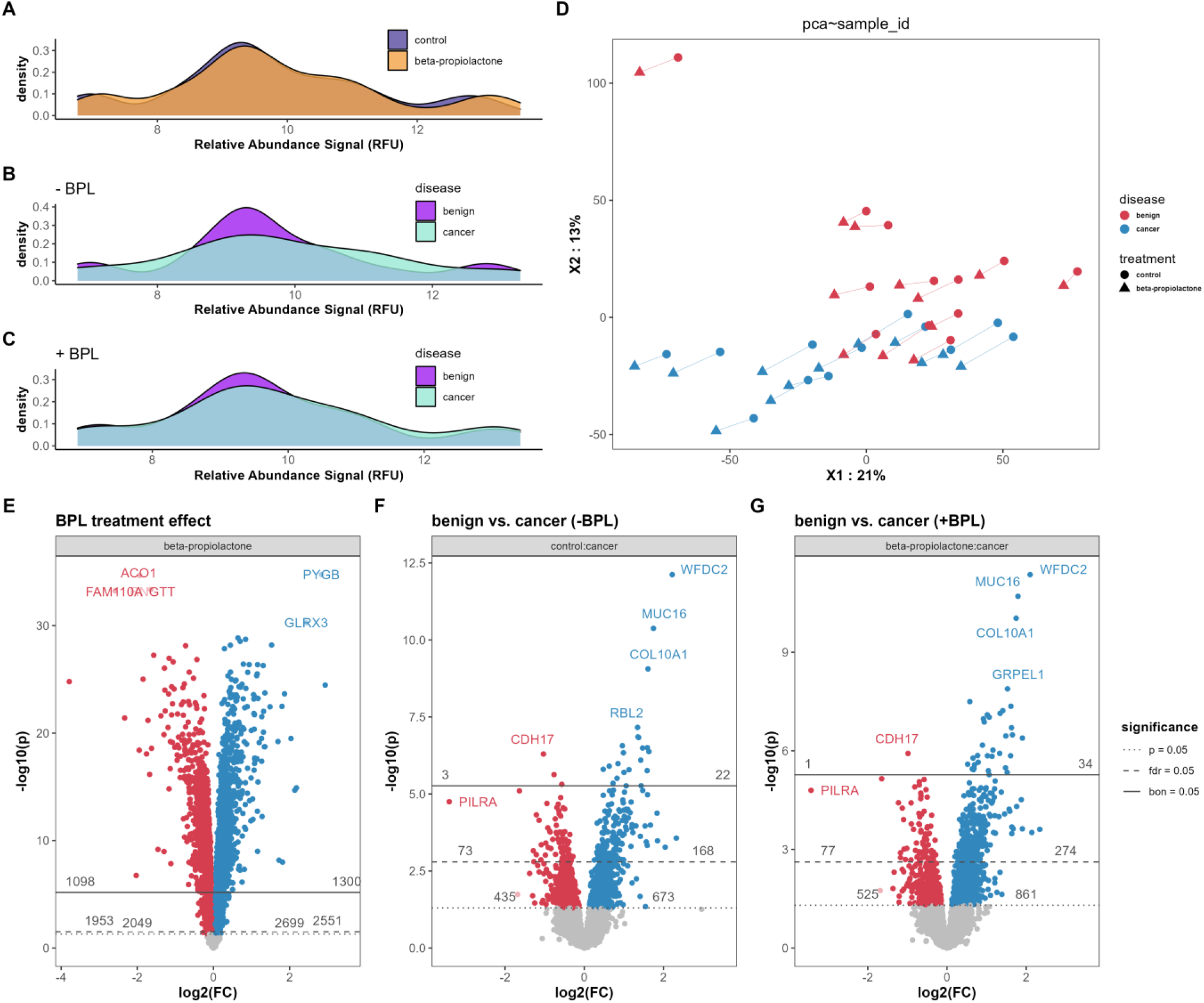
**A-C** Feature density plots showing the distribution of measured protein RFUs. **A** Comparison of RFU distribution in BPL-treated (orange) and untreated samples (blue). **B+C** Comparison of RFU distribution in cancer (turquoise) and benign samples (purple), without (B) and with (C) BPL treatment. **D** Principal Component analysis. PCA with coloring based on disease state (benign = red, cancer = blue), shape indicates treatment (circle = untreated, triangle = BPL-treated), samples from same subjects are connected with a line. **E-G** Volcano plots of proteins with differentially plasma abundance. **E** Volcano plot showing differences in SomaScan measurements between BPL treated and untreated samples, assesed by limma. Mapped are 1098 proteins with lower measurements (red) and 1300 proteins with higher measurements (blue) after BPL treatment (bon. adj. p < 0.05). **F** Volcano plot of distribution of differentially abundant proteins in cancer measured without BPL treatment. Three proteins are downregulated (red), while 22 proteins are upregulated (blue). **G** Differentially abundant proteins in benign vs. cancer measured after BPL sample treatment. One protein is significantly downregulated (red) and 34 proteins are upregulated (blue) in cancer (bon. adj. p < 0.05).

Differential expression analysis using Bayesian moderated t-testing as implemented by limma [11], supported these assumptions (Figure1 E-G). This analysis revealed that 2398 of ∼7000 (34%) targets showed significantly (Bonferroni corrected p < 0.05) different measurements after BPL treatment independent of disease state (Figure 1E). Of these, 1098 proteins were measured with reduced abundance, while 1300 were measured at higher abundance, when comparing BPL treatment to mock controls. The proteins most affected by BPL treatment were ACO1 (SomaScan SeqId: 20054-289, PYGB (SomaScan SeqId: 24414-3), FAM110A (SomaScan SeqId: 22371-46), RNGTT (SomaScan SeqId: 20135-85) and GLRX3 (SomaScan SeqId: 16596-25). Figure 1F and G show the differential expression of proteins in samples from subjects with benign tumors versus patients with ovarian cancer without (Figure 1F) and with BPL treatment (Figure 1G), respectively. As apparent from the volcano plots, the same three proteins (WFDC2, MUC16 and COL10A1) emerge to strongly differentiate ovarian cancer from benign plasma samples irrespective of BPL treatment.

From the RFU distribution of the most BPL treatment-affected proteins in differential expression analysis, it is apparent that despite significant altered measurements, samples from benign tumor patients and samples from ovarian cancer patients are affected to a comparable extent and the relative ratio between benign and cancer samples was maintained, preserving detectability of group differences. This is exemplified in Figure 2 A-E for the five proteins with most significantly changed abundance between ovarian cancer and benign samples in our data set, as detected by differential expression analysis. Figure 2 F-J shows the proteins for which the assays were most affected by BPL treatment.

**Figure 2:**
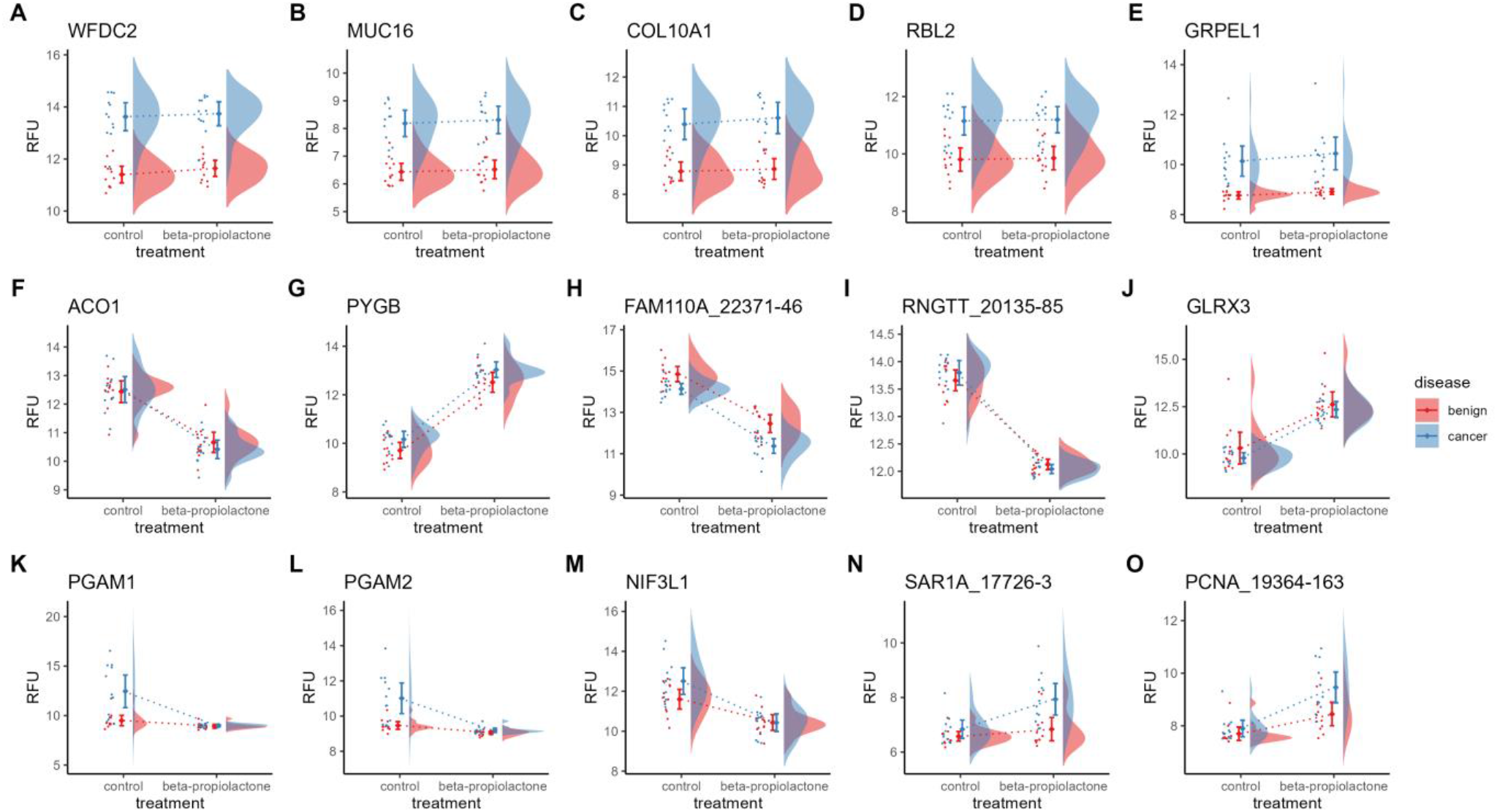
**A-E** Raincloud plots for protein features with most significant differences between benign and cancer samples. **F - J** Raincloud plots of proteins showing highest diversion of RFUs upon BPL treatment. **K – O** Raincloud plots of proteins showing the highest interaction effects between cancer and BPL treatment.

ACO1, FAM110A and RNGTT were measured with significantly lower abundance following BPL treatment, while PYGB and GLRX3 were measured with higher abundance. The ratios between protein levels in cancer versus benign samples, however, did not change significantly. On the other hand, a small number of protein targets presented with significantly changed ratios, exemplified by the five strongest affected targets in Figure 2 K-O.

To understand how BPL treatment impacts the effect size between measured protein abundances in benign versus cancer samples, both with and without BPL treatment globally, we calculated Cohen’s d [12] for all protein features. Figure 3 A represents the corresponding graphical analysis for all effect sizes. The median effect size between protein abundances in benign vs. cancer samples in BPL treated samples was d = 0.449, and d = 0.436 in untreated control comparisons. The resulting effect size difference Δd = 0.013 appeared significant (p < 0.05). Breaking this down to small (d < 0.2), medium (0.2 < d < 0.8) and large effect sizes (d > 0.8), it became apparent that primarily large effect sizes were impacted by the BPL-mediated increase (Figure 3 B-D). Globally, the benign vs. cancer effect sizes for every assayed protein, both with and without BPL treatment, showed an excellent correlation (ρ > 0.9) (Figure 3 E - G).

**Figure 3:**
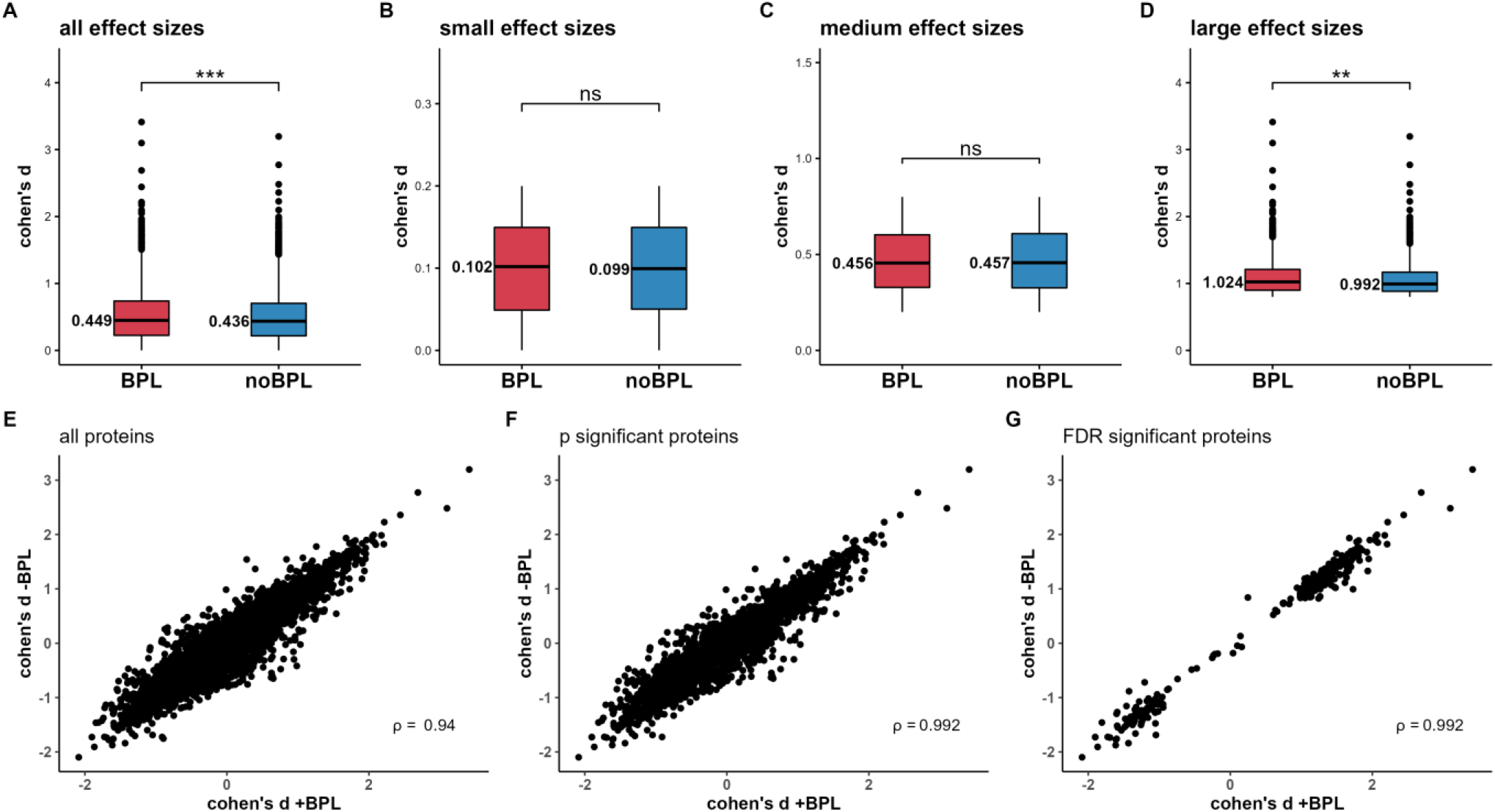
Analysis of changes in Cohen’s d effect sizes between protein targets upon BPL treatment. **A** Cohen’s d over all protein features. **B** Comparison of effect sizes between benign and ovarian cancer samples for features showing small effect sizes (d < 0.2), medium effect sizes (d > 0.2 and d < 0.8) **C** and large effect sizes (d > 0.8) **D. E-G** Correlation of Cohen’s d effect sizes between cancer and benign sample protein level means with and without BPL treatment. Pearson correlation of all (**E**, ρ = 0.94), p significant (**F**, ρ = 0.98) and FDR significant proteins (**G**, ρ = 0.98).

As sample- and assay-specific effects are lost during calculation of Cohen’s d, we were also interested in individual interaction effects and to that end investigated the interaction coefficients of a two-factor (treatment*disease) limma-driven linear model. Figure 4 shows a contrastogram providing an overview of significantly changed protein levels (FDR < 0.05) in the contrasts investigated. In Figure 4, C1 and C2 show the differences in measured protein abundance within either benign (C1) or cancer (C2) samples, representing the overall effect of BPL on the assay. A large percentage of proteins show altered measurements (44.14% and 54.57%), indicating that BPL changes the measurements of protein expression substantially. The impact on detection of differential protein expression between benign and cancer samples in BPL-treated and -untreated samples is indicated by C3 and C4, respectively. As a general trend, the number of altered proteins between cancer and control plasma samples increased upon BPL treatment. In the untreated samples, abundance of 73 proteins appeared decreased and of 168 proteins increased in cancer samples (Figure 4, C4, green arrows), while the numbers increased in BPL-treated samples to 77 and 274 (Figure 4, C3, yellow arrows), respectively. A total number of 42 (0.6%) assays showed significant interaction effects (C5; 27 positive, 15 negative) (Supp. File 1). The observed interaction effects ranged from -2.897 to 1.15 with a mean of 0.015.

**Figure 4:**
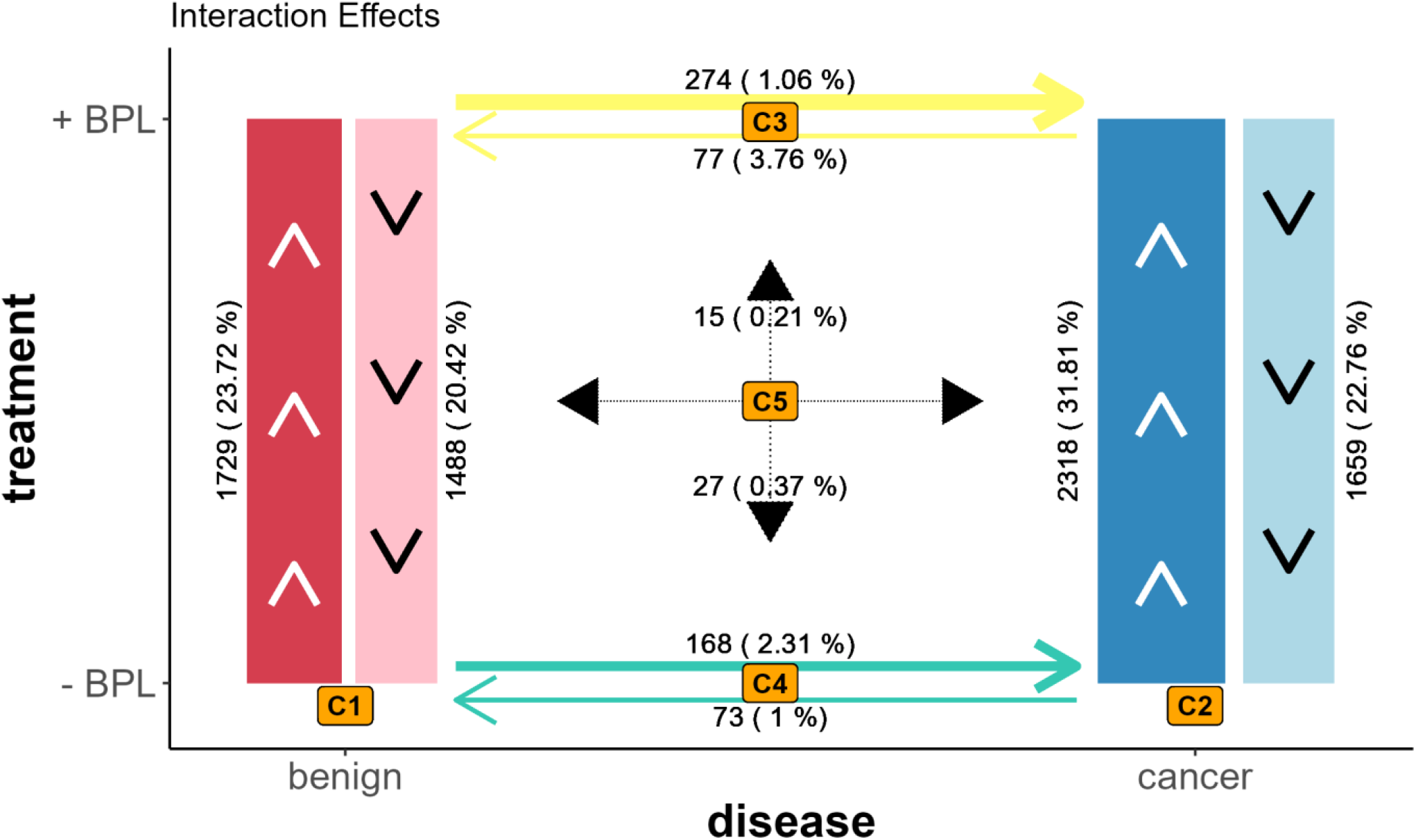
Contrastogram. Graph shows numbers of significantly altered protein levels in different contrasts of two-factor limma-based analysis (FDR < 0.05). Line thickness is proportional to number of proteins identified as differentially expressed. Arrow direction indicates the condition with higher expression. **Red** (C1: ∼disease/treatment; benign:BPL) Number of altered protein level measurements upon BPL treatment in benign samples only. Levels of 1729 proteins were measured increased and 1488 decreased. **Blue** (C2: ∼disease/treatment; cancer:BPL) Numbers of altered protein level measurements upon BPL treatment in cancer samples only. Levels of 2318 proteins were measured increased and 1659 decreased. **Yellow** (C3: ∼treatment/disease; BPL:cancer) Numbers of altered protein levels between benign and cancer samples in BPL-treated samples. Here, 274 proteins were measured with higher levels and 77 with lower levels. **Green** (C4: ∼treatment/disease; noBPL:cancer) Numbers of altered protein levels between benign and cancer samples in untreated samples. Here, 168 proteins were measured with higher levels and 73 proteins with lower levels. **Black** (C5: ∼treatment*disease; BPL:cancer) Interaction effects of BPL treatment and disease state on measured protein levels. Fifteen proteins show increased levels while 27 proteins show lower levels.

We investigated possible reasons for the enhanced and decreased signal levels observed upon BPL treatment. To that end, as BPL is known to be most reactive towards specific amino acid residues (especially cysteine, methionine and histidine) [6], we analyzed whether the amino acid composition of strongly affected proteins reflected this and calculated the relative abundance of every amino acid in the top 100 significantly altered measurements. The relative amino acid frequencies for proteins with increased measurements ranged from 0.9 to 1.14 with significantly (one-sample t test, p < 0.05)) increased phenylalanine, isoleucine, valine and tyrosine and decreased glutamic acid, proline, arginine and serine frequencies. Highest changes were observed for isoleucine and proline with frequencies >10% deviating from expected values (Figure 5A). The relative amino acid frequencies for proteins with decreased abundance measurements ranged between 0.89 and 1.11, with a significant increase of alanine and methionine frequency of 11% and a significant decrease of serine and threonine frequency (Figure 5B)(Supp. File 1). Even though the identification of ovarian cancer biomarkers was not the main goal, the present study also revealed a number of novel potential candidates that were not detectable by our previous analysis based on smaller assay panels [13], notably COL10A1 (Figure 1F and G). Intriguingly, interrogation of the in PRECOG database [10] revealed that COL10A1 gene expression is strongly associated with a short survival of ovarian cancer (z-score 5.40, corresponding to a p-value of 4.2×10^−6^), emphasizing the potential relevance of this finding. Our analysis also identified proteins clinically used as plasma biomarkers for ovarian cancer, i.e., WFDC2 and MUC16 [13]–[17], further validating the SomaScan technology for biomarker discovery.

**Figure 5:**
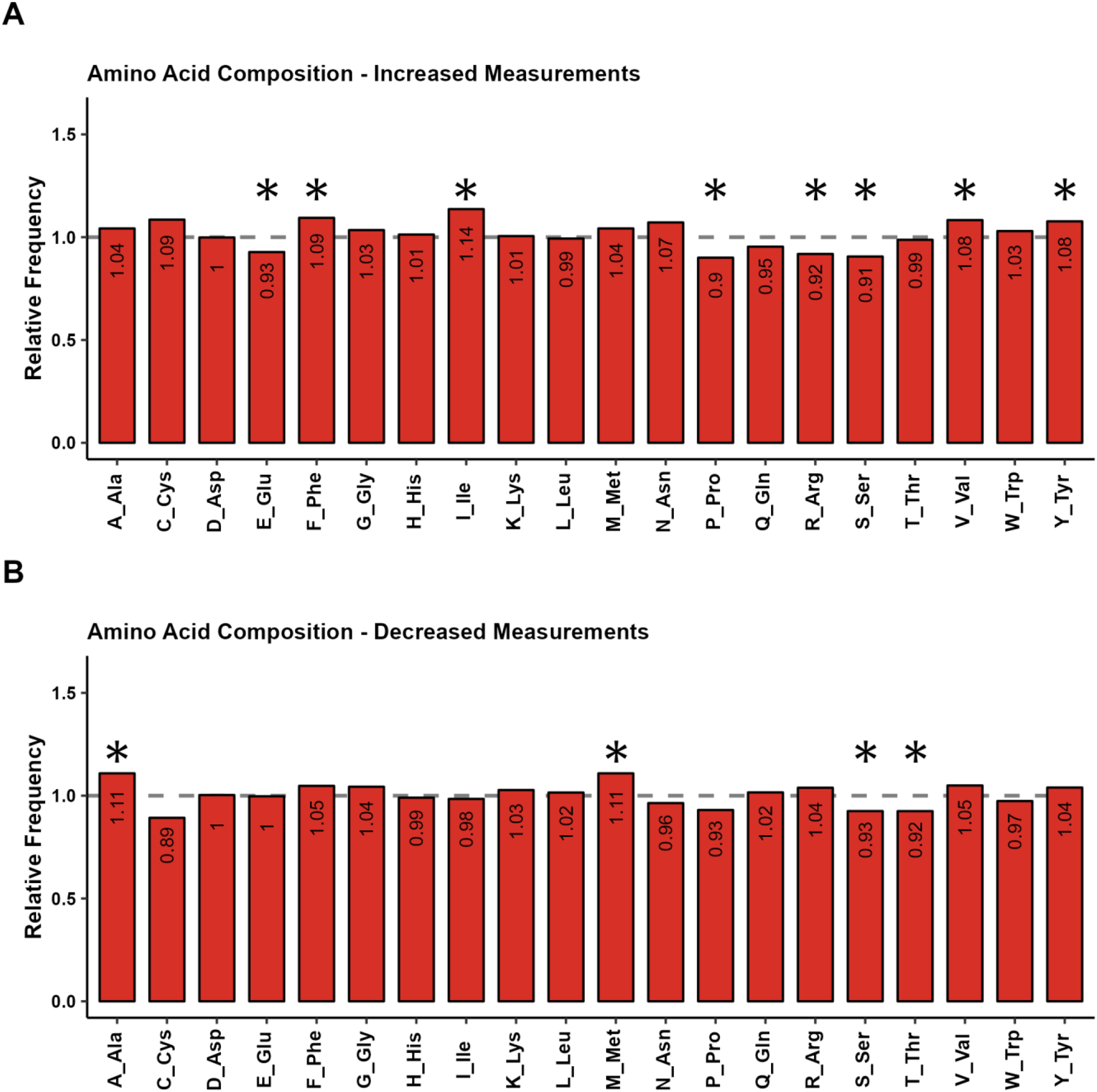
Amino acid compositions of proteins with increased RFU values (**A**) and decreased RFU values (**B**) after BPL treatment. Amino acid composition shows relative frequencies of each amino acid in the enhanced or impaired assays. Dashed line representing the expected frequency with regard to all proteins in the SomaScan panel normalized to 1. Asterisk indicates statistical significance (one-sample t test, p < 0.05) (Supp. File 1).

## Discussion

In this study, plasma samples from ovarian cancer patients were compared to samples from patients with benign tumors with the primary goal to evaluate detectability of proteins after BPL treatment using the SomaScan platform. Our results show that a substantial proportion of protein assays was significantly impacted by treatment with BPL prior to SomaScan analysis. Approximately one third of assays showed altered detection, indicating a major impact of BPL treatment on the analysis (Figure 1E & Figure 4 C1+C2). Despite this obvious effect of the BPL treatment, the crucial aspect for the assessment of suitability of SomaScan assays in its context lies in determining whether it remains possible to analyze differential protein expression in e.g. different study groups. Our results show that the group differences in protein expression measurement is largely preserved following BPL treatment (Figure 1F-G, Figure 4 C3-C4). The protein assays strongly affected by the treatment showed an equivalent alteration in both treatment groups, indicating that differential protein expression may still be measured even under the influence of BPL (Figure 2 F-J). The ratio between protein levels in benign and cancer samples was altered in only a minute number of assays (0.6%), as indicated by significant interaction effects of treatment and disease state in linear modeling (Figure 2 K-O, Figure 4 C5). Differential expression measurements for these proteins are compromised by the BPL treatment. A slight increase in significantly changed protein levels between benign and cancer samples was observed upon BPL treatment. When investigating Cohen’s d effect sizes of the measured differences between every assay from benign and cancer samples, a minor but significant increase was observed in the BPL-treated samples (Figure 3A), which is also apparent in interaction effects observed using linear modeling and further reflected in the differential expression analysis, showing a higher number of significant group differences (Figure 4 C3 vs C4). This phenomenon may ultimately increase false positive hits and thus effort required for downstream validation studies. On a global scale, the differences in the means of protein levels from benign and cancer samples were maintained upon BPL-treatment, as indicated by the high correlation between pre- and post-treatment effect sizes (Figure 3 E-G).

As BPL is known to be most reactive with the amino acids cysteine, methionine and histidine, while essentially non-reactive towards asparagine, glutamine and tryptophan [6], we hypothesized that the protein assays most affected by the treatment might target proteins with an amino acid composition reflecting this. The relative amino acid frequencies of proteins with enhanced measurements showed on average a 14% increase of isoleucine and a 10% decrease of proline compared to the expected values. Proteins with decreased measurements showed an 11% higher frequency of alanine and methionine, as well as an 11% decrease of cysteine frequency on average. While isoleucine, proline and alanine are not known to be reactive to the reagent, BPL causes alkylation and acylation of cysteine and alkylation of methionine [6]. Modifications of these residues as well as conformational changes in the respective proteins potentially caused by such modification, may influence epitope availability and consequently the binding affinity of the aptamers used in SomaScan. She et al. reported a correlation of altered protein conformation and folding with the positions of modified amino acid residues [8]. Also, BPL-modified residues may potentially mask SOMAmer-targeted epitopes by steric hindrance [8]. To fully explain BPL induced changes in the measured signal intensity, additional, individual analysis of the respective targets and knowledge of the epitopes the targeting SOMAmers rely on are needed and are beyond the scope of the present study.

BPL’s use in virus inactivation is primarily rationalized by its alkylation and acetylation of guanosine and adenosine, as well as induction of DNA cross-linking [18]. The SomaScan technology is based on SOMAmers as protein binders. These molecules are single stranded deoxyoligonucleotides enhanced with protein-like functional groups, mimicking amino acid side chains that are selected from large random libraries using SELEX [9]. SOMAmers are thus potentially modified by BPL, which might alter binding affinity to their target proteins. BPL in aqueous solutions undergoes continuous hydrolysis with a half life of 16– 20h at 4°C [1]. The samples used in this study were incubated for 72h at 4°C prior to SomaScan analysis, followed by weeks of storage and transport at -20°C and on dry ice, respectively. As it was previously recommended to allow four days for complete hydrolysis of BPL [1], the presence of residual BPL in the samples cannot entirely be ruled out, yet appears unlikely.

Although the focus of this study is not on the differences in plasma protein levels in samples from patients with ovarian cancer and benign tumors, the results of this group comparison shall be briefly discussed here. The top markers for ovarian cancer in comparison to benign patients identified in our study are WFDC2, MUC16 and COL10A1. The former two, also known as HE4 and CA125, are well recognized plasma biomarkers for ovarian cancer and were evaluated in numerous studies [13]–[17] while COL10A1 has not been proposed as an ovarian cancer biomarker. This result is of great potential interest, as the PRECOG dataset [10] indicates a highly significant association of COL10A1 gene expression with a short survival in ovarian cancer, which is consistent with the reported association of COL10A1 with other cancer entities [19]–[21].

In conclusion, the BPL-treatment of plasma samples does not appear to significantly and globally impact the detection of differential protein expression using SomaScan. A small subset of protein targets is, however, significantly affected, showing corresponding interaction effects. Based on the available data, we are unable to derive mechanistic insight into this altered detection behavior. We finally conclude that SomaScan appears to be a viable tool for the proteomic analysis of blood-derived samples even in the context of special handling needs in high-risk biosafety environments and expect this finding to extend to affinity proteomics platforms in general.

## Data Availability

All data produced in the present study are available upon reasonable request to the authors

## Acknowledgments

The authors are indebted to Prof. Dr. Stephan Becker, Institute for Virology, Philipps-Universität Marburg (Germany) for sharing the BPL inactivation protocol. The authors thank SomaLogic Operating Co., Inc. (Boulder, Colorado) for, in the context of the study presented here, providing access to the SomaScan platform free of charge.

